# Incidence and Determinants of loss to follow-up of HIV pre-exposure prophylaxis among key and priority population groups in Addis Ababa health centers, Ethiopia retrospective cohort study

**DOI:** 10.1101/2023.09.28.23296311

**Authors:** Addisu Aragaw, Fentaw Taddse

**Affiliations:** Department of public health, GAMBY Medical and Business college; PO Box 106 (C-1035); Addis Ababa Ethiopia; Department of Epidemiology, Wollo University; Wollo Ethiopia.

## Abstract

**Background:** - ‘loss of follow-up’ is a significant public health concern globally. Retention in preventive care among at-risk key and priority population groups is critical for the successful prevention of HIV acquisition. Loss to follow-up of individuals has countless negative impacts on HIV prevention outcomes. There is, however, limited information about the incidence and predictors of loss to follow-up in the study area. Thus, this study aimed to determine the incidence rate and predictors of loss to follow-up among Prep users.

**Method:** - A retrospective cohort study was undertaken using 240 PrEP users between May 2020, and May 26, 2022, at Addis Ababa selected health centers. All eligible clients who fulfilled the inclusion criteria were included in study.

**Results and discussion:** A total of 240 participants with a median age of 32.0 years (interquartile range [IQR]: 27.0 to 40.0) had a median time since initiation of PrEP 21.2 months (IQR: 6.5 to 22.1). Almost half 122 (50.8%) were married, single 58 (24.2%), divorced 10 (4.2%), and widowed 50 (20.8%). One third were female sex workers (33.3%), and the rest (66.7%) were sero-discordant couples. Most of the users were from ART clinics (60.8%) and (39.2%) were from PMTCT clinics Being male is associated with 1.77 times in risk of getting lost than female (ARR=1.77, 95% CI =1.12-2.79). those who had no adherence counseling was associated with 1.86 times in the chance of LTFU as compared to those who had have adherence counseling (ARR=1.863, 95% CI =1.184, 2.930, P-value=0.003). 86 (35.8%) experienced loss to follow-up and the overall incidence rate of loss to follow-up was 7.3 (95% CI: 4.3-12.6) per 100 person-years of observation (PYs). PrEP adherence strategies should be developed and designed as a holistic approach, acknowledging the contextual factors of key population groups.

**Conclusion:** - About 35.8% clients became LTFU of PrEP with overall incidence of 7.3(4.3-12.6) per 100 pys observation. Research preparedness involving key and priority population groups should be strengthened for HIV prevention intervention evaluations in Ethiopia.

## INTRODUCTION

Health is among the seventeen sustainable development goals (SDGs) whose promise is to end the epidemic of AIDS, tuberculosis, malaria among others by the year 2030. (1) Globally, by 2017, the number of people living with AIDS (PLHIA) were estimated to be between 31.1 to 43.9 million with those newly infected (all ages) being approximately 1.4 to 2.4 million. (2)

Human immunodeficiency virus infection and acquired immune deficiency syndrome (HIV/AIDS) continue to be a major global public health issue. By the end of 2018, an estimated 37.9 million individuals worldwide were living with HIV/AIDS. (3, 4) HIV remains a global health crisis. In 2020, there were: 37.9 million people living with HIV, including 10.2 million who were not on treatment: 1.5 million New HIV infections and 680 000 AIDS-related deaths. (5) Overall, key populations and their sexual partners accounted for 65% of HIV infections. worldwide in 2020 and 93% of infections outside of sub-Saharan Africa. (5) However, not all population subcategories face the same risk of acquiring HIV/AIDS. (4)

Globally and in Africa specifically, female sex workers (FSWs) are at an extraordinarily high risk of contracting HIV.(6) Globally Sex workers are accounted 11%, People who inject drugs 9%,,Transgender women 2% Remaining Population 35% Clients of sex workers and sex partners of all key populations 20% (5) and In sub-Sahara Africa Sex workers are 12% People who inject drugs are 1% Gay men and other men who have sex with men 6% Transgender Women 1% Clients of sex workers and sex partners of all key populations 19%,Remaining population are 61%.(7) An estimated 29.3% of FSWs were living with HIV/AIDS in sub-Saharan Africa in 2014.(8) Meanwhile, research has indicated that sex workers and clients of sex workers and other sexual partners account for 3% and 19% of new yearly HIV infections, respectively.

Oral pre-exposure prophylaxis (PrEP) is a novel biomedical HIV prevention intervention recommended for use by the World Health Organization (WHO) among key populations including FSWs. (16) To avert this ongoing new HIV infection, a combination HIV prevention approach including PrEP as an additional component is crucial Oral tenofovir disoproxil fumarate (TDF) is the currently recommended PrEP regimen for women medication which is recommended to be taken daily. It offers a locus of control for women to be in charge of their HIV prevention choices. (17) Considering this, PrEP is endorsed in the national comprehensive HIV Prevention, care and treatment guideline and HIV prevention roadmap (2018–2020). Currently, the PrEP services have been provided in public health facilities and DICs for FSWs and HIV negative sero-discordant partners. (15)

Globally they make up to 9% new HIV infections. They are 13 times more at risk of HIV as compared to the general population due to an increased likelihood of being economically vulnerable, unable to negotiate consistent condom use and experiencing violence, criminalization and marginalization. (9) and make up a small proportion of the general population, but they are at extremely high risk of HIV infection. (10)

During the last two decades, the Ethiopia responded vigorously to curb the epidemic through a multi-sectorial approach involving all stakeholders, mobilizing resources and the community at large (11). This may result in failure to achieve the desired programmatic objectives in reducing HIV incidence due to high dropout rates among clients initiated on PrEP who may end up being exposed to HIV acquisition risk. In order to achieve desired programmatic objectives; this study sought to describe possible contributors to PrEP lost follow up as well as factors which may affect continued use of PrEP. Counseling services provided for HIV negative clients to remain negative and provision of PrEP uses as one strategy to reduce new infections and HIV acquisition Currently, characteristics and factors of Pre-exposure prophylaxis (PrEp) is part of HIV testing and antiretroviral medicine based strategies among other combinations prevention programs towards attainments of UNAIDS targets of three 95 accessing tailored with three million individuals alone targeted for PrEP.(12)

The targeted populations include female sex workers, sero discordant couples and their sexual partners in high prevalence sites supported by PEPFAR. Ethiopia started rolling out PrEP in late 2019, targeting female sex workers and HIV discordant couples. For the initial phase, 15,400 female sex workers and 4,762, couples in discordant relationships were targeted. By December 2019, the number of people screened for PrEP services were 4,128, of which 1589, were found eligible, 971 were initiated while 601 (61.9%) declined. (13) The current HIV epidemic in the Federal Democratic Republic of Ethiopia is low intensity, mixed epidemic kind with important non uniformity across geographic areas and population groups. The national HIV prevalence is calculated to be 0.9%, urban areas are a lot more affected than rural areas (2.9% versus 0.4%) whereas females are double as affected as male (1.2% versus 0.6%). Out of the eleven regional states and town administrations, seven have HIV prevalence of 1% and above. Key and priority populations (KP & PP) are disproportionally affected compared with the national average: 23% among feminine Sex staff (FSW), 5.1 % discordant couples.

In HIV negative partners of sero-discordant couples: There is no reliable data source/s on the level of discordance. However, more than 60% of adult PLHIV and 80% of sexually active PLHIV are currently married and report relatively little extra marital risk behaviors. Roughly twothirds of these have sero-discordant sexual partners. (12) Discordant couples have the highest risk of acquiring HIV. From the total HIV positive couples in Addis Ababa,5.1 % of them were found to be discordant. (14) In Ethiopia, there are still new infections annually largely moving the Key population and priority population groups. From WHO’s recommendation, Pre-Exposure prophylaxis (PrEP) of HIV is one in all the interventions that facilitate countries to cut back new HIV infection. (6)

Ethiopia may not reach its UNAIDS goals to end ADIS by 2030 without specifically addressing among key population and as far as my knowledge concerns there is no literature in Ethiopian on assessing incidence and determinants of lost follow up of pre-exposure prophylaxis users among key population. Our study therefore assessed incidence and determinants of lost follow up of pre-exposure prophylaxis users among key population in Addis Ababa.

Ethiopia has committed itself reaching the goal of 95-95-95 targets towards the management and control of HIV adopted the use of PrEP as among the most recent robust and effective HIV preventions interventions (15).

Carrying out this study is important to determine the lost follow up factors of HIV preexposure prophylaxis (PrEP) and the factors associated with PrEP and their adherence level in each follow up visit among key population groups. Therefore, the national program requires further understanding on what is driving this challenge in order to come up with programmatic interventions to improve PrEP continuation rates.

Despite the rapid expansion of antiretroviral therapy services, ‘loss to follow-up’ is a significant public health concern globally. Loss to follow-up of individuals from pre –exposure follow up has a countless negative impact on the incidences for HIV and its prevention.

Loss to follow-up is a serious public health concern throughout the world. In Ethiopia, there is limited information regarding incidence and predictors of loss to follow-up among key population groups on pre-exposure prophylaxis users. It is worth it to scale up PrEP service nationwide to contribute to the reduction of new HIV infections.

The study is designed to provide baseline data for program planners, decision-makers, and pre-exposure prophylaxis implementers at a different level. The study provides baseline information for governmental and non -governmental organization that works in collaboration with the federal ministry of health’s area of HIV/AIDS particularly on pre-exposure prophylaxis’s study provides information regarding the current factors associated with pre-exposure loss to follow-up and the existing gap.

## METHODS

### Study Design and Recruitment

The study was conducted in Addis Ababa city administration health centers is the capital of Ethiopia. The national central statistical agency that carries out national census projections put the population of Addis Ababa to be 3,273, 000 (CSA, 2015) based on this figure, the population of Addis Ababa accounts for 3.6% of the national population and 18% of the urban population in Ethiopia. It has shown an annual rate of population growth of 2.1 %. (9) In Addis Ababa city administration, there are different governmental, private and non-governmental health facilities which provide HIV comprehensive services. The city administration has 11 sub cities and 99 health centers and among these five health centers were selected for the study being conducted. The study was conducted from May 1, 2022 to June 30, 2022.Retrospective cohort study design was employed to determine the incidence and determinants of loss to follow-up (LTFU) of HIV pre-Exposure prophylaxis.

#### Source population

All clients of pre-exposure prophylaxis who were initiated on PrEP at Addis Ababa Health centers. *Study population*: All clients of pre-exposure prophylaxis who were initiated on PrEP from May 30, 2020, to May 26, 2022, at Addis Ababa selected health centers.

The sample size was determined by using the EPI INFO version 7.2.3.1 using the cohort study design formula. From the study, the proportion of among FSW exposed with loss to follow up was 49. %, and among non-exposed who have lost follow up of none FSW were 68.9% (45).

Considering 95% confidence interval, 80% power. Finally, by using one to two ratios of the exposed to non-exposed ratio (1:2), then the final total sample size was found to be 240 (80 exposed and 160 non exposed groups).

Exposed group Study participants who were female sex worker during study period was considered as exposed group and as a Non-Exposed group Study participants who were not female sex worker.

#### Sampling Procedure

A simple random sampling technique was employed. Among 11 sub-cities in Addis Ababa five health centers were selected from the selected sub-cities. Then a total sample of 240 was distributed to the five health centers based on the proportion of PrEP takers in each health center.

**Figure 1.**
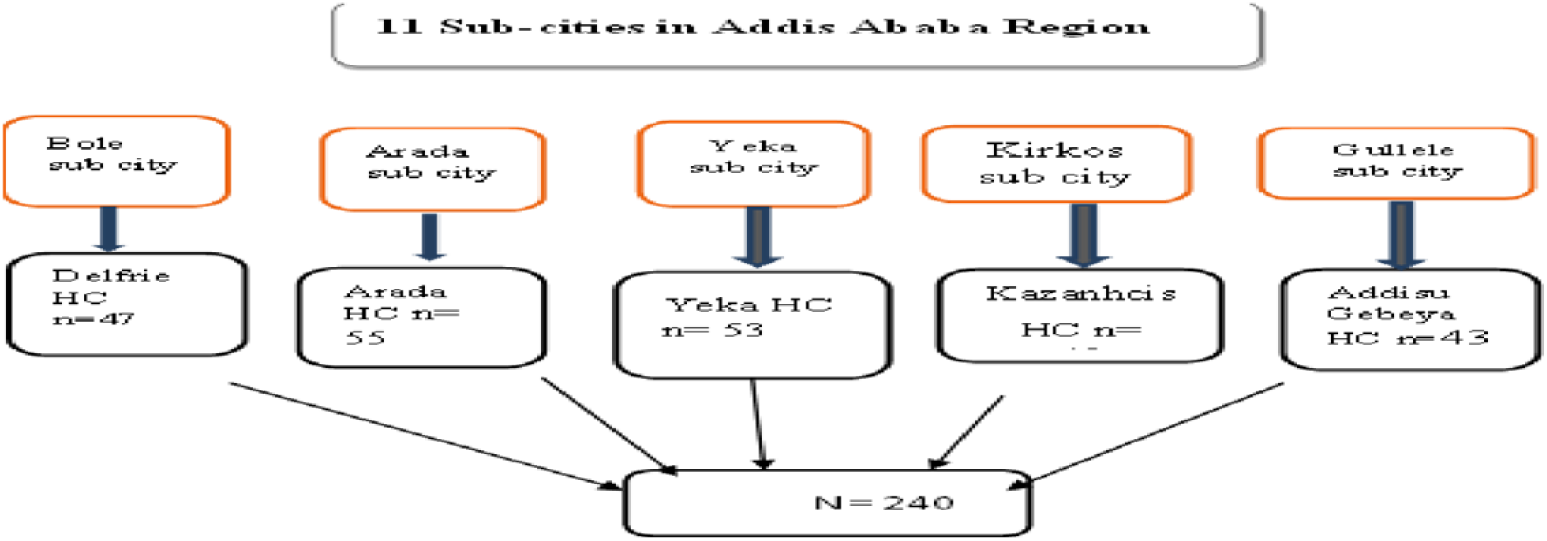
Schematic presentation for sampling techniques

Inclusion critrrea All selected participants who started pre –Exposure prophylaxis in between December,2020 and First December,2022 was included based on the record review. Clintes excluded from analysis, (had an incorrect medical record number (n=16), did not meet criteria for PrEP (n=11), who had in complete information(n=13)), the study excluded clients who were transferred to other health facilities.

As Outcome/dependent Variable Loss to follow up of pre-exposure prophylaxis which is dichotomies as 1 if “Yes” and 0 “No”.

##### Independent Variable

Socio demographic or Socio-economic factors: sex, age, marital status, phone functionality, region/address and level of education, Baseline clinical and laboratory information: - Baseline HBsAg Test result, Pregnancy Status, Syndrome STI Screening and STI Diagnosis result, Pre exposure follows up status: - Transfer out, Transfer in, Died, Lost, and Missed Dose, Pre exposure prophylaxis follow up information: - adherences status, drug side effect, HIV retest status, pregnancy status, use, type family planning, - Female Sex Workers and Sero Discordant Couples

##### Operational definitions

*Lost to follow up (LTFU)* if they were late by >90 days for their scheduled appointment date (16). Pre-Exposure Prophylaxis (PrEP): - Defined as the use of antiretroviral drugs by HIV-negative people, before potential exposure to prevent the acquisition of HIV (11).

Key populations: - are populations who are at higher risk for HIV irrespective of the epidemic type or local context and who face social and legal challenges that increase their vulnerability or describe a category of populations of special interest in the HIV epidemic (17).

Adherence: - Is defined as those who are missing less than 4 doses have good adherences and missing 5 plus dose are poor adherences (18).

Female sex workers: - are defined as women who regularly or occasionally exchange sex for money in drinking establishments, night clubs, local drink houses, “chat” and ‘’shisha’’ houses, “on the street”, around military and refugee camps, construction sites, trade routes, red light districts, and at their homes

##### Ethical consideration

The program evaluation was approved by Addis Ababa Public Health Research and Emergency Management Directorate review board and the school of GAMBY medical and Business College’s ethical review committee. Human research protection procedures were determined to be research, but investigators did not interact with human subjects or have access to identifiable data or specimens for research purposes. Individual consent to use clients’ secondary data was waived by the IRBs.

#### Statistical Analysis

The data were edited, cleaned, coded, and entered into Epi-info window version 7.2.3.1 and analyzed using SPSS windows version 20.

Descriptive statistics i.e., frequencies were run to check for outliers. Data were summarized using frequencies and percentages for categorical covariates and numerical were summarized with mean ± standard deviation (SD) or medians with interquartile range (IQR) values as required. Baseline characteristics of participants who had follow-up and those who were lost-to-follow-up were compared by using Chi-square tests (for categorical variables) or independent sample t-tests (for continuous variables).

The association between the dependent variables and the outcome of pre-exposure follow-up was analyzed using the robust Poisson regression model. Univariate robust regression was run at a 25% level of significance to screen our potential significance of independent variables. The multivariable robust Poisson regression model was performed using significant independent variables from univariate robust Poisson regression.

To measure the presence and strength of associations between the predictor and outcome variable relative risk (RR), a p-value and 95% CI were used. From the final model, variables with a P-value ≤ 0.05 were considered as significantly associated with the outcome of loss to follow-up pre-exposure prophylaxis.

The overall fitness of the model (model adequacy) was assessed by using the goodness of fit test and omnibus test.

## RESULTS

Among key and priority population groups who were taking PrEP registered for the program from 2019 –2022 data of 240 were retrieved from PrEP registration and client’s card for the study. At the beginning of PrEP started the median age was 32.0 years (interquartile range [IQR]: 27.0 to 40.0). Out of 240 participants followed during the study period, almost two-third 154 (64.2%) were females and the remaining 86 (35.8%) were males. Almost half 122 (50.8%) were married, single 58 (24.2%), divorced 10 (4.2%), and widowed 50 (20.8%). Regarding educational status participants with no formal education 68 (28.3%), primary education 97 (40.4%), secondary education 56 (23.3%) college and above 19 (7.9%). More than half of 124 (51.7%) of them had functional cell phone and others had no functional cell phone 116 (48.3%) and most of 220 (91.2%) lived in the study area, Addis Ababa and 20(8.8% lived out of Addis Ababa (Table 1).

**Table 1.** baseline socio demographic characteristics of Individuals (n=240)

| Characteristics | Categories | Frequency (n) | Percentage (%) |
| --- | --- | --- | --- |
| **Sex** | Male | 86 | 35.8 |
|  | Female | 154 | 64.2 |
| **Age** | 21-30 | 82 | 34.2 |
|  | 31-40 | 103 | 42.9 |
|  | 41-50 | 48 | 20.0 |
|  | 51-60 | 7 | 2.9 |
| **Marital status** | Single | 58 | 24.2 |
|  | Married | 122 | 50.8 |
|  | Divorced | 50 | 20.8 |
|  | Widowed | 10 | 4.2 |
| **Living Address** | Within Addis Ababa | 220 | 91.2 |
|  | Out of Addis Ababa | 20 | 8.8 |

**Table 2.** HIV Risk Classification and PrEP Service Delivery Point of Individuals Started PrEP (n=240)

| Characteristics | Categories | Frequency (n) | Percentage (%) |
| --- | --- | --- | --- |
| **PrEP provision site** | ART | 146 | 60.8 |
|  | PMTCT | 94 | 39.2 |
| **HIV risk classification** | Female sex workers | 80 | 33.3 |
|  | None female sex workers | 160 | 66.7 |

**Table 3.** Baseline clinical and laboratory information of Individuals Started PrEP (n=240)

| Characteristics | Categories | Frequency (n) | Percentage (%) |
| --- | --- | --- | --- |
| **Screened for Hepatitis** | Yes | 121 | 50.4 |
|  | No | 119 | 49.6 |
| **Hepatitis testing result** | Not done | 127 | 52.9 |
|  | Negative | 113 | 47.1 |
| **Pregnancy testing result** | Positive | 19 | 11.1 |
|  | Negative | 153 | 88.9 |
| **screened for TB** | Yes | 202 | 84.2 |
|  | No | 38 | 15.8 |
| **Screened for STI** | Yes | 201 | 83.8 |
|  | No | 39 | 16.3 |
| **Diagnosed with STI** | Yes | 78 | 32.5 |
|  | No | 153 | 63.8 |
| **STI developed by type** | Vaginal Discharge | 33 | 16.3 |
|  | Ureteral discharge | 17 | 8.3 |

majority of them 211 (87.9%) had first follow-up visit and didn’t, have 29 (12.1%). more than half 125 (52.1%) had a Second follow-up visit the retested no (47.9%) had no, and only 87 (36.3%) had a third follow-up visit but 153 (63.8) of them didn’t have third follow up visit. All clients were assessed for any signs and symptoms of acute HIV infection 240 (100%), HIV retest status, and negative 152 (63.3%). No sero-conversion, not done 88 (36.7%), among female Pregnancy follow-up status and Negative was 96 (40%) among them 18 (7.5%) were pregnant, and the rest 49 (20.4%) didn’t know their pregnancy status during the follow-up visit. Number, of missed tablets 48 (20) % of them missed more than five tablets, 67 (27.9%) had missed ≤4 tablets, and more than half 125 (52.1%) didn’t know the amount of missed tablet.165 (68.8%) had got adherence counseling during follow-up time but 75 (31.1%) did not, from them Risk redaction provided 192(80%), among them 39 (36.3%) good Adherence 192 (63.8%) had poor adherences. Only116 (48.3%) used family planning methods among these, 19 (7.9%) used condom, 10 (4.2%) OCP, injectable 67(27.9%), IUCD and others 20 (8.4%). Only 13% Developed drug side effects from them 19(7.9%) Abdominal pain, Vomiting and Fever 20 (8.4%) (Table 4).

**Table 4.**
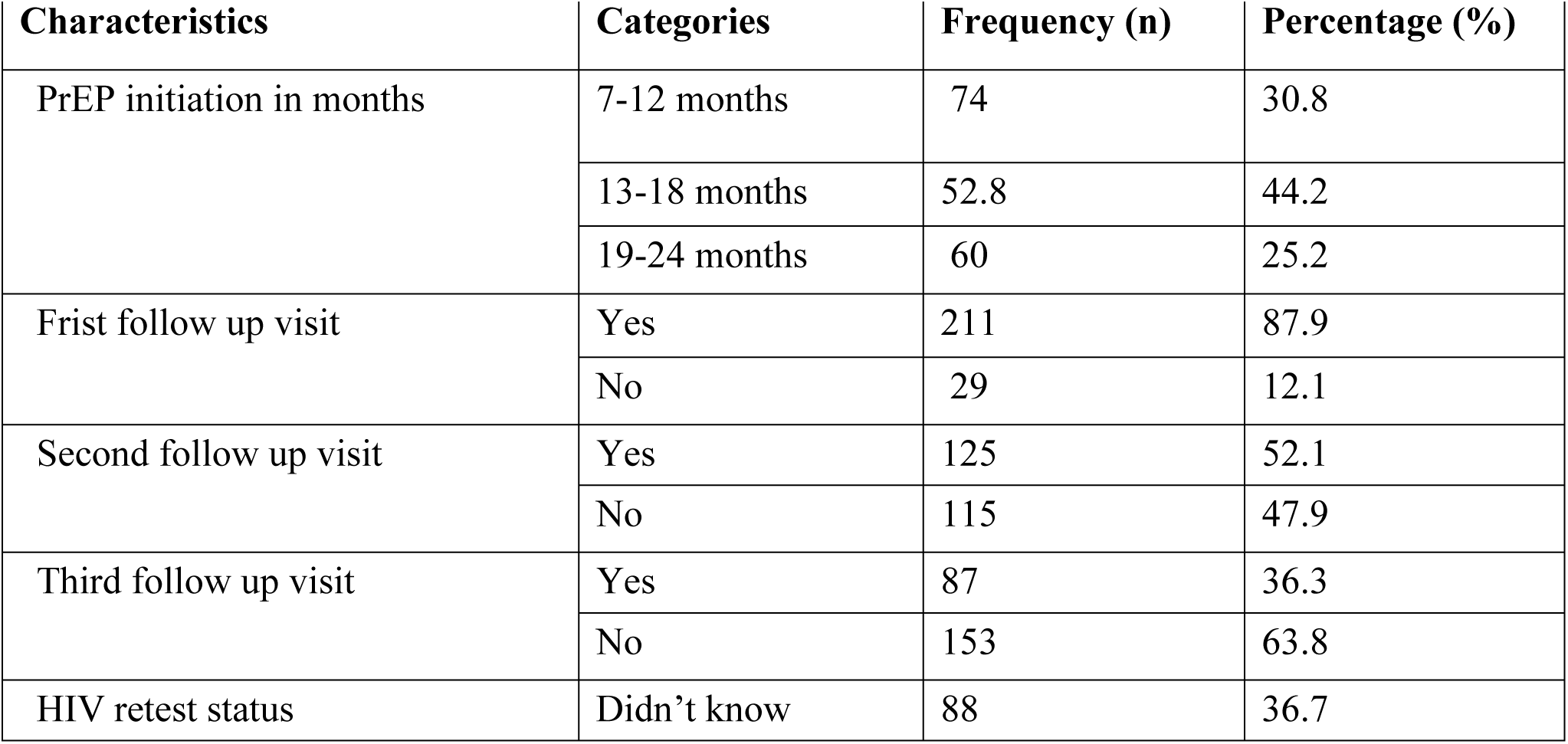
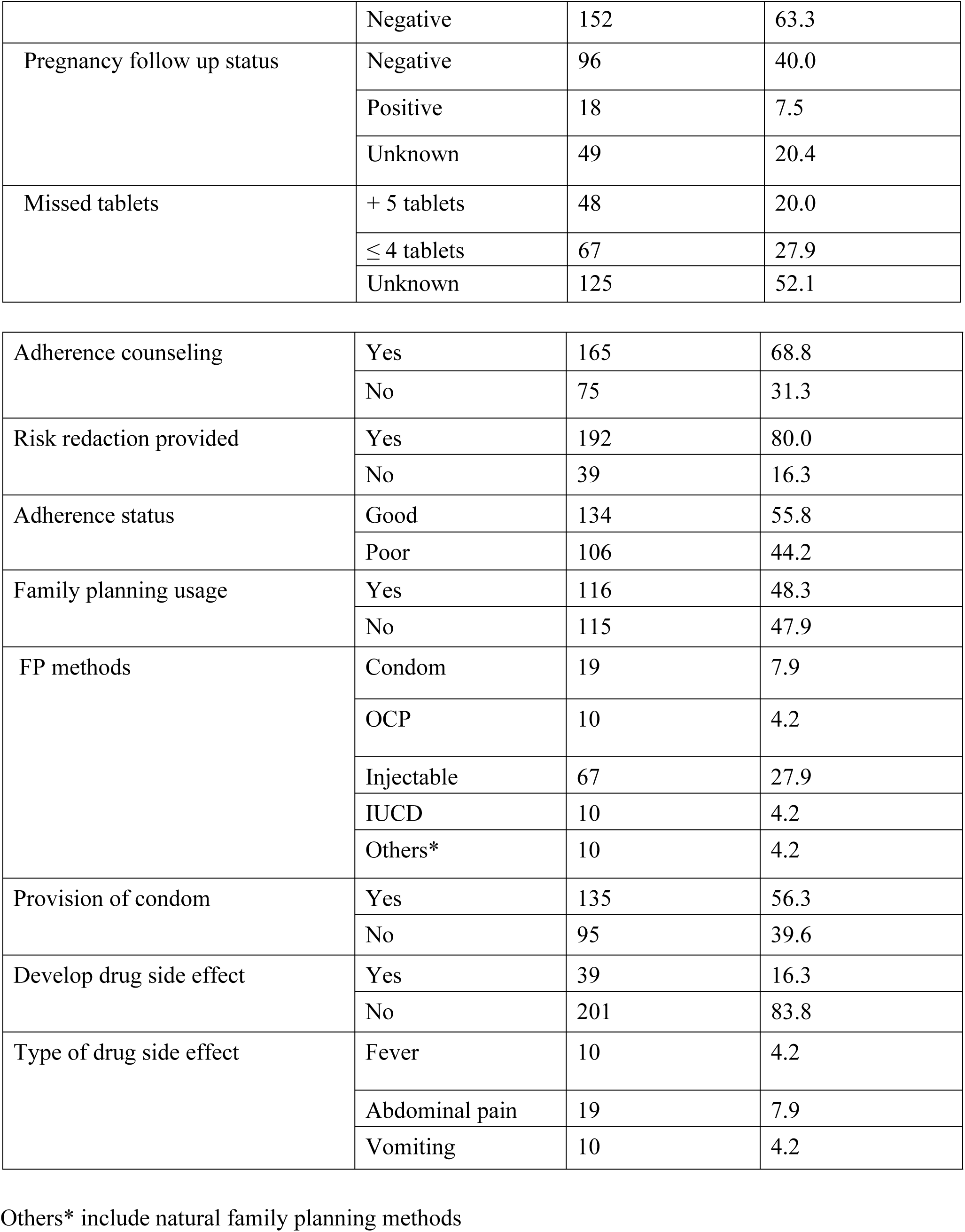
PrEP initiation and clinical follow up assessment of individuals started PrEP (n=240)

| Characteristics | Categories | Frequency (n) | Percentage (%) |
| --- | --- | --- | --- |
| PrEP initiation in months | 7-12 months | 74 | 30.8 |
|  | 13-18 months | 52.8 | 44.2 |
|  | 19-24 months | 60 | 25.2 |
| Frist follow up visit | Yes | 211 | 87.9 |
|  | No | 29 | 12.1 |
| Second follow up visit | Yes | 125 | 52.1 |
|  | No | 115 | 47.9 |
| Third follow up visit | Yes | 87 | 36.3 |
|  | No | 153 | 63.8 |
| HIV retest status | Didn't know | 88 | 36.7 |
|  | Negative | 152 | 63.3 |
| Pregnancy follow up status | Negative | 96 | 40.0 |
|  | Positive | 18 | 7.5 |
|  | Unknown | 49 | 20.4 |
| Missed tablets | + 5 tablets | 48 | 20.0 |
|  | ≤ 4 tablets | 67 | 27.9 |
|  | Unknown | 125 | 52.1 |
| Adherence counseling | Yes | 165 | 68.8 |
|  | No | 75 | 31.3 |
| Risk reduction provided | Yes | 192 | 80.0 |
|  | No | 39 | 16.3 |
| Adherence status | Good | 134 | 55.8 |
|  | Poor | 106 | 44.2 |
| Family planning usage | Yes | 116 | 48.3 |
|  | No | 115 | 47.9 |
| FP methods | Condom | 19 | 7.9 |
|  | OCP | 10 | 4.2 |
|  | Injectable | 67 | 27.9 |
|  | IUCD | 10 | 4.2 |
|  | Others* | 10 | 4.2 |
| Provision of condom | Yes | 135 | 56.3 |
|  | No | 95 | 39.6 |
| Develop drug side effect | Yes | 39 | 16.3 |
|  | No | 201 | 83.8 |
| Type of drug side effect | Fever | 10 | 4.2 |
|  | Abdominal pain | 19 | 7.9 |
|  | Vomiting | 10 | 4.2 |

Others\* include natural family planning methods

The incidence rate of LTFU of PrEP were followed for a minimum of 7 and maximum of 23 months of the follow-up period. The total follows up period was 346 person-months (288.7 person years) of observation, and the median follow-up period was 21.1 (IQR: 6.5 to 22.1). At the end of the follow-up period, a total of 86 (35.8%) patients experienced loss to follow-up. Hence, the overall incidence of loss to follow-up was estimated to be 7.3 (95% CI: 4.3-12.6) per 100 person years of observation.

### Analyses of robust Poisson regression with outcome of LTFU of PrEP

Univariate analysis at a 25% level of confidence was run to select the variables to fit into the multivariable analysis robust Poisson regression model and from which Sex, Age, Phone, HIV risk category, Hepatitis test result, Pregnancy status, TB result, Screened for STI, first and third follow up visit, pregnancy follows up status, family planning methods usage, provision of adherence counseling, were found to be significant.

In the final model, after adjusting for other covariates being male is associated with a 1.77 times higher risk of getting lost as compared to being female (ARR=1.770, 95% CI =1.120, 2.797). In addition, the risk of getting lost from PrEP follow-up youngest (21–30) years was 0.406 times lower than those aged (51–60) years (ARR=0.406, 95% CI =0.223, 0.738, P-value=0.003). similarly, those in the age of (31–40) years was associated with a 1.77 times risk of loss compared with (51–60) years (ARR= 0.391, 0.641 Vs. 1.77 95% CI).

Those clients who were knowing their hepatitis B results were associated with 0.29 times lower than who didn’t know their results (ARR=0.29, 95% CI =0.20-0.42). Clients who didn’t Screen for STI were 8.53 times higher than those who were screened for STI (ARR=8.53, 95% CI=4.10-17.74).

Those who had a first follow-up visit according to their follow-up time were associated with 18.1% lower chances for lost than as compared to those who didn’t have a first follow-up visit (ARR=1.81, 95% CI=1.211-2.72). In the addition, those who had a didn’t have third follow-up visit according to their follow-up time were associated with 1.08 times for lost than as compared to those who have a third follow-up visit (ARR=1.08, 95% CI=1.27, 2.59). Among female PrEP users, those who didn’t know their pregnancy status during each follow-up visit were 4.28 times higher in the risk of LTFU compared to who did know about their pregnancy (ARR=4.28, 95% CI=3.04, 6.01).

Those clients who had no adherence counseling were associated with 1.86% times higher in the risk of LTFU as compared to those who had adherence counseling (ARR=1.86, 95% CI =1.18, 2.93). Those who didn’t used family planning methods had 2.29 times higher in the chances of LTFU of PrEP than those who used any family planning methods (ARR=2.29, 95% CI =1.39, 3.78) table 5.

**Table 5.**
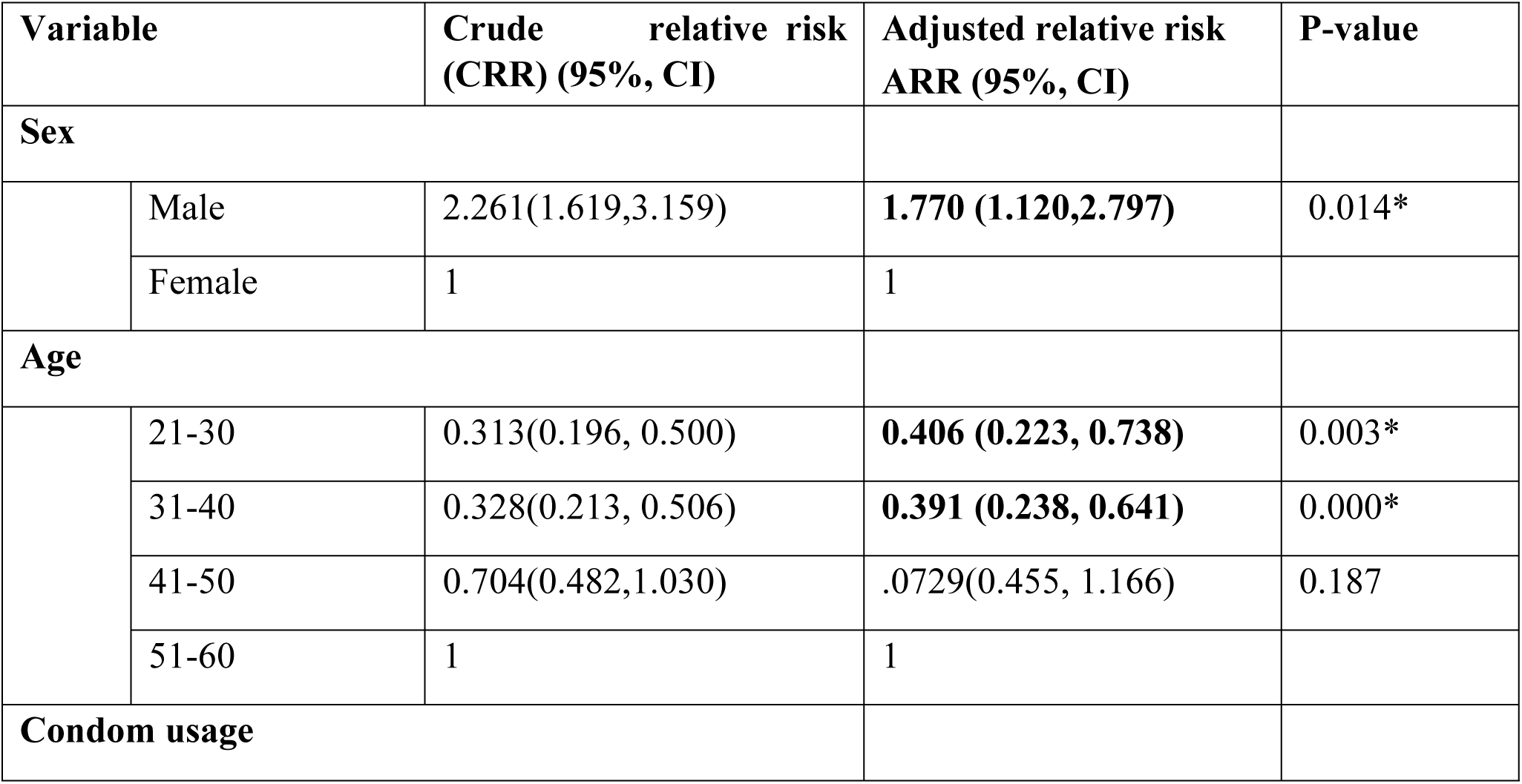
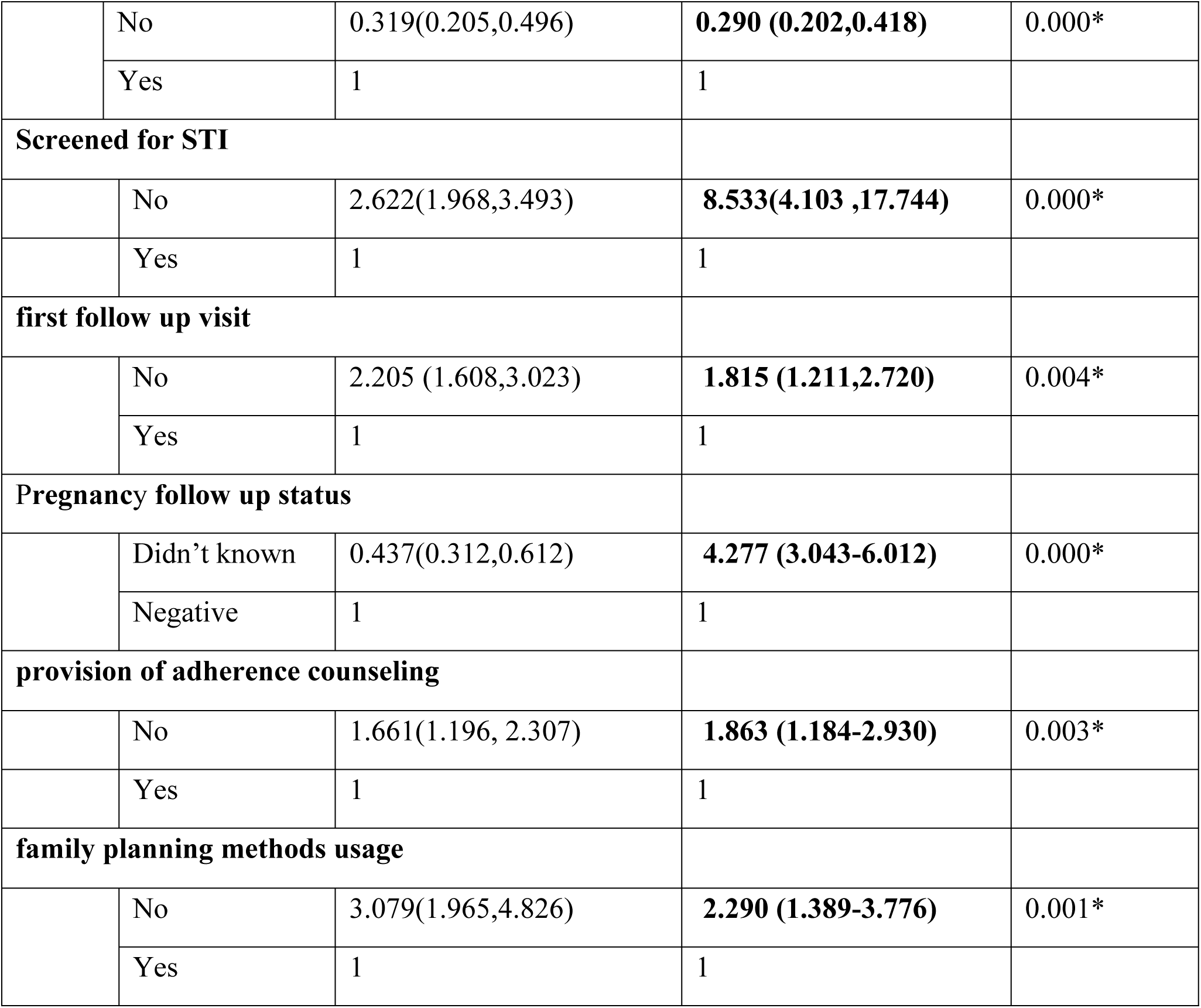
Risk factor associated with lost follow up of Pre exposure users (n=240).

| Variable |  | Crude relative risk (CRR) (95%, CI) | Adjusted relative risk ARR (95%, CI) | P-value |
| --- | --- | --- | --- | --- |
| **Sex** |  |  |  |  |
|  | Male | 2.261(1.619,3.159) | **1.770 (1.120,2.797)** | 0.014* |
|  | Female | 1 | 1 |  |
| **Age** |  |  |  |  |
|  | 21-30 | 0.313(0.196, 0.500) | **0.406 (0.223, 0.738)** | 0.003* |
|  | 31-40 | 0.328(0.213, 0.506) | **0.391 (0.238, 0.641)** | 0.000* |
|  | 41-50 | 0.704(0.482,1.030) | .0729(0.455, 1.166) | 0.187 |
|  | 51-60 | 1 | 1 |  |
| **Condom usage** |  |  |  |  |
|  | No | 0.319(0.205,0.496) | **0.290 (0.202,0.418)** | 0.000* |
|  | Yes | 1 | 1 |  |
| **Screened for STI** |  |  |  |  |
|  | No | 2.622(1.968,3.493) | **8.533(4.103 ,17.744)** | 0.000* |
|  | Yes | 1 | 1 |  |
| **first follow up visit** |  |  |  |  |
|  | No | 2.205 (1.608,3.023) | **1.815 (1.211,2.720)** | 0.004* |
|  | Yes | 1 | 1 |  |
| **Pregnancy follow up status** |  |  |  |  |
|  | Didn't known | 0.437(0.312,0.612) | **4.277 (3.043-6.012)** | 0.000* |
|  | Negative | 1 | 1 |  |
| **provision of adherence counseling** |  |  |  |  |
|  | No | 1.661(1.196, 2.307) | **1.863 (1.184-2.930)** | 0.003* |
|  | Yes | 1 | 1 |  |
| **family planning methods usage** |  |  |  |  |
|  | No | 3.079(1.965,4.826) | **2.290 (1.389-3.776)** | 0.001* |
|  | Yes | 1 | 1 |  |

## DISCUSSION

Loss to follow-up is the major difficulty and it complicates the evaluation of HIV prevention and intervention programs. This study assessed the incidence and its predictors of loss to follow-up among key and priority population groups who started PrEP. In this study, 86 (35.8%) experienced loss to follow-up and the overall incidence rate of loss to follow-up was 7.3 (95% CI: 4.3-12.6) per 100 person-years of observation (PYs). In this study, we document a higher LTFU rate than previously reported in the PrEP period (35.8 vs. 23.9 per 100PY) (19).

In the study found that gender was significantly associated with lost follow up of PrEP. The incident rate for male PrEP user is 1.77 times the incidence rate for females. High adherences is especially important among male, as compared to women using PrEP, to achieve high efficacy of PrEP because of differences in perception and health seeking behavior (20).

Approach based on their gender during initiation of PrEP and during their follow up time based on their needs, Providers’ suggestion and support of PrEP use have also been suggested as effective solutions to improve retention in PrEP care in a variety of diverse situations (21, 22). This study shows that PrEP patients are at high risk of loss to follow-up at the early stage of PrEP initiation was 1.815 times LTFU the incidence who didn’t have frist follow up visit). Probability of retention in PrEP care among patients who are in preexposure prophylaxis. At the time of stage of PrEP initiation could be promising to prevent loss to follow-up among those likely to discontinue. Targeted interventions are needed to focus on populations currently underserved by existing services to improve retention in PrEP care (23).

Likewise, the incident rate in the age of 21-30 and 31-40 years is 0.406 and 0.39 times the incident rate for 51-60 years of age groups remaining other variables are constant respctivly. The possible reason might be that the younger age group could be more mobile than the older one and may face the fear of discrimination and stigma. The other possible reasons might be immaturity in analytical thinking and particular challenges associated with young age(24).

Provision of adherence counselling had a statistically significant association with the incidence rate of LTFU. Those clients who hadn’t have adherence counselling were 1.66 times at higher risk of loss to follow-up compared to those with adherence counseling. “The possible explanation might be that those health providers provision adherence counselling levels may have different skills practice, counseling problems that affect client’s adherence initially and during follow up time which further affect their retention in PrEP user.”

Using contraception methods with clients were significantly associated with LTFU of PrEP. in the study showed that high LTFU to PrEP has been found among PrEP user who didn’t use contraception methods. We can therefore see a link between lost follow up and contraceptive use and therefore intensified contraceptive counseling and screening before PrEP initiation and during PrEP use may improve PrEP adherence among users (25).

From the results, LTFU to PrEP has also been found to be higher among who do not use condoms; this is alarming since PrEP is being promoted as a method of HIV prevention that should be used in combination, or as a back-up method that one can use when she fails to use condoms. In this case counselors need to emphasize the need adhere to PrEP well if from their interactions they find that the use of condoms is challenging for the client. The the higher LTFU to PrEP among users who don’t use condoms could have been due to poor HIV risk perception and misunderstandings about the duration of action of PrEP to protect against HIV.

The health belief model (26) explains that people will not change their health behaviors unless they believe that they are at risk; in this case, those who do not think that they are at risk of acquiring HIV are unlikely to use a condom or take PrEP. This furthermore underscores the need for interventions to improve the evaluation of risk perception among PrEP users. Qualitative data may better explain higher LTFU among lower condom use.

In the finding, there was a total of 35.8% lost follow up of PrEP which is similar studies done in USA was 36% (27). A study Nigeria LTFU was 66.7% and in this study 77% due to adherences problem (28). poor adherences to PrEP, history of STI and younger age was associated with lost follow up of PrEP (28). another study in Uganda LTFU of PrEP in female sex worker was 42% (29) which is similarly to 39.6% in this study. Given from the findings, and evidence in the literature, PrEP clients had highest LTFU happened in first follow up visit 32% (30) and in this study, it was 76.7%.

### Study Strengths and Limitations

While this study provides information on lost follow-up of clients and supports future development and improvement of retention among pre-exposure prophylaxis users a number of limitations were encountered through the health center in the study design.

Patient record availability was limited, due to the recent implementation of a PrEP in late 2019 and lack of registration system.

Due to the nature of the secondary source of data, some missed variables like status of mental illness, stigma and discrimination, distance from the health facility, and economic status, might be predictors. In addition to these, charts with an incomplete record of 20% and transferred outpatients were excluded; this might lead to underestimating or overestimating the incidence rate of loss to follow-up. Information bias, specifically misclassification of exposure status, may have been present if the client’s chart did not answer questions.

Missing data impacted a number of variables evaluated in this study; this may lead to selection bias. As strength the it Fills gaps in research and become Baseline for further studies.

## CONCLUSIONS

Barriers to PrEP adherence such as use of contraception need to be addressed for successful PrEP implementation to improve adherence going forward. There is need for service care providers to reinforce positive behaviors such as use of other HIV prevention measures such as condoms devotedly when there is a break from taking PrEP. Intensified contraceptive counseling and screening may improve PrEP adherence among users.

Therefore, PrEP adherence strategies should be developed and designed as a holistic approach, acknowledging the contextual factors of key and priority populace groups. centered interventions thinking of socio-demographic and individual preferences have the capability to assure the highest adherence stage.

### Recommendations

Based on the above findings, federal MOH of Ethiopia should adopt and incorporate WHO guidelines for these key and priority population in HIV prevention should be implemented at each level of health centers. the Findings suggest that comprehensive strategies are warranted to improve PrEP continuum of care outcomes in key and priority populations.

Multi-modal strategies are most impactful for retention of PrEP after initiation PrEP adherence strategies should be developed and designed as a holistic approach, each health care providers and concerned bodies should Use the current protocol prepend by of MOH Ethiopia for those key and priority population in HIV prevention and care like, Client centered and Targeted interventions approach is needed,Gender based approach during initiation of PrEP, Targeted interventions focus at first initiation,Provision of adherence counselling,Intensified contraceptive counseling and screening, Emphasize on provision and of use of condom.

A qualitative study should be applied to obtain information from LTFU patients themselves by tracing them and the health service quality and satisfaction level of patients need to be addressed.

Furthermore, a prospective cohort study needs to be conducted and a qualitative study should be applied to obtain information from LTFU patients themselves by tracing them and the health service quality and satisfaction level of patients need to be addressed.

## Data Availability

Before publication, I can submit the required to make fully available and without restriction all data underlying the findings based PLOS Data Policy page for detailed information on this policy.

## Acknowledgment

I amplify my true appreciation to my advisor Fentaw Tadese who made a difference in me beginning from proposal writing up to point-by-point investigation and introduction of my thesis work.

My extraordinary obliged to go to all health Center authorities for permitting me to urge the essential data and all the medical Professional for their dynamic participation all through the information collection period. And goes to my families who are continuously making a difference on the side me up to my completion of the work.

## References

1. Grimsrud A, Barnabas RV, Ehrenkranz P. Evidence for scale up: the differentiated care research agenda. J Int AIDS Soc. 2017; 20:1–6.

2. FMOH, National JSS REPORT, 2021.

3. UNAIDS. Get on the Fast Track. Geneva, 2016.

4. WHO: Consolidated guidelines on the use of antiretroviral drugs for treating and preventing HIV infection: recommendations for a public health approach – 2nd ed., 2016.

5. Grimsrud A, Bygrave H, Doherty M, et al. Reimagining HIV service delivery: the role of differentiated care from prevention to suppression. JIAS. 2016, 19:21484.

6. International AIDS Society. Differentiated Care for HIV: A Decision Framework for ART Delivery, 2016..

7. Fast-Track – ending the AIDS epidemic by 2030. Geneva: UNAIDS; 2014 (http://www.unaids.org/en/resources/documents/2014/JC2686_WAD2014report, accessed 1 November 2015)..

8. Fox MP, Rosen S. Retention of Adult Patient on Antiretroviral Therapy in Low- and Middle-Income Countries: Systematic Review and Meta-analysis 2008-2013. J Acquir Immune Defic Syndr. 2015; 69(1):98–108.

9. Abnet G, Dawit B, Imam M, Tsion L, Yonas A. City profile of Addis Ababa. EiABC-Ethiopian Institute of Architecture Building Construction and City Development. 2017:5–7.

10. DATA U. UNAIDS reference. 2019.

11. Implementation manual for pre -exposure prophylaxis (PrEP) of HIV infection. In: EMOH, editor. 2019.

12. UNAIDS. On the fast track to end acquired immunity defiance diseases (AIDS). 2016.

13. Boender TS, Sigaloff KC, McMahon JH et al. Long-term Virological Outcomes of FirstLine Antiretroviral Therapy for HIV-1 in Low- and Middle-Income Countries: A Systematic Review and Meta-analysis. Clin Infect Dis. 2015;61(9):1453–61..

14. Central Statistical Agency (CSA) [Ethiopia] and ICF International. Ethiopia Demographic and Health Survey: HIV Report. Addis Ababa, Ethiopia, and Rockville, Maryland, USA. 2016.

15. Anyona MO. Determinants of condition of pre-exposure prophylaxis. 2019.

16. Elizabeth W., Wahome. Grahama N. Et.al. PrEP uptake and adherence in relation to HIV1 incidence among Kenyan men who have sex with men. The lancet. September 2020.

17. WHO. Guideline on when to start Antiretroviral Therapy and on Pre-Exposure Prophylaxis for HIV (2015) accessed on 2017.

18. Baral S, Beyrer C, Muessig K, et al. Burden of HIV among female sex workers in lowincome and middle-income countries: a systematic review and meta-analysis. Lancet Infect Dis. 2012; 12:538–49.

19. Pillay D, Stankevitz K, Lanham M, Ridgeway K, Murire M, Briedenhann E, et al. Factors influencing uptake, continuation, and discontinuation of oral PrEP among clients at sex worker and MSM facilities in South Africa. PLoS One. 2020;15:e0228620.

20. Sheth AN, Rolle CP, Gandhi M. HIV pre-exposure prophylaxis for women. Journal of Virus Eradication. 2016;2(3):149. https://www.ncbi.nlm.nih.gov/pmc/articles/PMC4967966/ Accessed on 7/28/2020.

21. Kenison TC, Badenhop B, Safo S. Unlocking HIV preexposure prophylaxis delivery: Examining the role of HIV providers in pre-exposure prophylaxis care. AIDS Patient Care STDs 2020; 34:251–258.

22. BrantAR, Dhillon P, Hull S, et al. IntegratingHIV pre-exposure prophylaxis into family planning care: A RE-AIM framework evaluation. AIDS Patient Care STDs 2020; 34:259–266.

23. Jun T, Madeline C, Montgomery W, et al. Loss to Follow-Up and Re-Engagement in HIV Pre-Exposure Prophylaxis Care in the United States, AIDS PATIENT CARE and STDs Volume 35, Number 7, 2021 @Mary Ann Liebert, Inc. DOI: 10.1089/apc.2021.0074 2013–2019. 24.

24. Mathewos A, Mekdes K, Haymanot N, Melkamu M, A. D. Predictors of Loss to Follow-Up among HIV-Infected Adults after Initiation of the First-Line Antiretroviral Therapy at Arba Minch General Hospital, Southern Ethiopia: A 5-Year Retrospective Cohort Study. Hindawi BioMed Research International Volume 2021, Article ID 8659372, 10.1155/2021/8659372. Received 8 June 2021; Revised 25 October 2021; Accepted 30 October 2021; Published 11 November 2021.

25. Nalukwago GK, Isunju JB, Muwonge T, Katairo T, Bunani N, Semitala F, et al. Adherence to oral HIV pre-exposure prophylaxis among female sex workers in Kampala, Uganda. Afri Health Sci. 2021;21(3). 1048–1058. 10.4314/ahs.v21i3.12.

26. Rosenstock IM. The health belief model and preventive health behavior. Health Education Monographs. 1974;2(4):354–86. 10.1177%2F109019817400200405 Accessed on 7/28/2020.

27. Douglas K, Kevin M, Victoria E, et al. Patterns and clinical consequences of discontinuing HIV preexposure prophylaxis during primary care Krakower D et al. Journal of the International AIDS Society, 22:e25250 http://onlinelibrary.wiley.com/doi/10.1002/jia2.25250/full | 10.1002/jia2.25250. 2019.

28. Adetunji O, Adewale S, M K. Pre-exposure prophylaxis in sero-discordant male partners of human immune deficiency virus (HIV) positive women desirous of natural conception – a clinical setting experience, Journal of AIDS and HIV Research. july,2013; Vol. 5(7), : 269–74.DOI 10.5897/JAHR2013.0252 ISSN 2141-2359 © 2013 Academic Journals http://www.academicjournals.org/JAHR

29. Joseph K, B J, Hadijja N, et al. Uptake and retention on HIV pre-exposure prophylaxis among key and priority populations in South-Central Uganda,Journal of the International AIDS Society, 23:e25588 http://onlinelibrary.wiley.com/doi/10.1002/jia2.25588/full | 10.1002/jia2.25588. 2020.

30. Jason B, Rupa R, Rachel B Lessons from a Decade of Voluntary Medical Male Circumcision Implementation and their Application to HIV Pre-Exposure Prophylaxis Scale Up. SAGE. 2018.

